# Outcomes of convalescent plasma with defined high- versus lower-neutralizing antibody titers against SARS-CoV-2 among hospitalized patients: CoronaVirus Inactivating Plasma (CoVIP), double-blind phase 2 study

**DOI:** 10.1101/2022.04.29.22274387

**Authors:** Luther A. Bartelt, Alena J. Markmann, Bridget Nelson, Jessica Keys, Heather Root, Heather I. Henderson, JoAnn Kuruc, Caroline Baker, D. Ryan Bhowmik, Yixuan J. Hou, Lakshmanane Premkumar, Caleb Cornaby, John L. Schmitz, Susan Weiss, Yara Park, Ralph Baric, Aravinda M. de Silva, Anne Lachiewicz, Sonia Napravnik, David van Duin, David M. Margolis

**Author notes:** Address correspondence to Luther A. Bartelt. Alternate correspondence to David M. Margolis.

## Abstract

**Background:** COVID-19 Convalescent Plasma (CCP) was an early and widely adopted putative therapy for severe COVID-19. Results from randomized control trials and observational studies have failed to demonstrate a clear therapeutic role for CCP for severe SARS-CoV-2 infection. Underlying these inconclusive findings is a broad heterogeneity in the concentrations of neutralizing antibodies (nAb) between different CCP donors. The present study was designed to evaluate nAb titer threshold for clinically effective CCP.

**Methods:** We conducted a double-blind, phase 2 study to evaluate the safety and effectiveness of nAb titer-defined CCP in adults admitted to an academic referral hospital. Patients positive on a SARS-CoV-2 nucleic acid amplification test and with symptoms for < 10 days were eligible. Participants received either CCP with nAb titers ≥1:160-1:640 (standard titer group) or >1:640 (high titer group) in addition to standard of care treatments. Adverse events were contrasted by CCP titer. The primary clinical outcome was time to hospital discharge, with mortality and respiratory support evaluated as secondary outcomes.

**Findings:** Between August 28 and December 4, 2020, 316 participants were screened, 55 received CCP, with 41 and 14 receiving standard versus high titer CCP, respectively. Participants were a median of 61 years of age (IQR 52-67), 36% women, 25% Black and 33% Hispanic. Severe adverse events (SAE) (≥ grade 3) occurred in 4 (29%) and 23 (56%) of participants in the high versus standard titer groups, respectively by day 28 (Risk Difference -0.28 [95% CI -0.56, 0.01]). There were no observed treatment-related AEs. By day 55, time to hospital discharge was shorter among participants receiving high versus standard titer, accounting for death as a competing event (hazard ratio 1.94 [95% CI 1.05, 3.58], Gray’s p=0.02).

**Interpretation:** In this phase 2 trial in a high-risk population of patients admitted for Covid-19, we found earlier time to hospital discharge and lower occurrences of life-threatening SAEs among participants receiving CCP with nAb titers >1:640 compared with participants receiving CCP with lower nAb titer CCP. Though limited by a small study size these findings support further study of high-nAb titer CCP defined as >1:640 in the treatment of COVID-19.

**Funding:** This clinical study was supported by the UNC Health Foundation and the North Carolina Policy Collaboratory at the University of North Carolina at Chapel Hill with funding from the North Carolina Coronavirus Relief Fund established and appropriated by the North Carolina General Assembly. The laboratory assays for neutralizing antibody titers and SARS-CoV-2 specific antibody-binding assays were partially supported by The NIH NCI/NIAID SeroNet Serocenter of Excellence Award U54 CA260543.

**Research in Context:** *Evidence before this study:* COVID-19 Convalescent Plasma (CCP) has emergency use authorization from the FDA for early treatment of COVID-19 in either outpatient or inpatient settings. Evidence supporting the use of CCP for severe COVID-19 is mixed and still emerging. One major limitation in interpreting published clinical trials and the clinical role of CCP is incomplete understanding of necessary neutralizing antibody (nAb) titer for clinically effective CCP. Observational studies suggest that higher antibody-content CCP is more effective than lower antibody-content CCP, or that very low antibody-content CCP is harmful. We searched PubMed articles published between February 1, 2020, and April 15, 2022, using the terms “COVID-19”, “convalescent plasma”, “SARS-CoV-2”, and “CCP” alone and in combination. Our search yielded 6,468 results which we filtered to 280 and 162 by selecting ‘Clinical Trial’ and ‘Randomized Controlled Trial’ article types, respectively. Among these, we identified 25 open-label or blinded efficacy or effectiveness studies in hospitalized patients that were relevant to our study. Preliminary reports show wide variability in the antibody content of CCP used in clinical trials, the assays used to define CCP antibody content, and the estimates of clinical outcomes following CCP therapy for hospitalized patients. Only one study deliberately infused CCP with nAb > 1:640. Post-hoc analyses of potent monoclonal antibody therapy in hospitalized patients in the UK showed survival benefit when monoclonal antibody was infused to patients who had not yet seroconverted by spike antibody ELISA, suggesting that if dosed appropriately, antibody-based therapies may have a role in improving outcomes of severe COVID-19.

*Added value of this study:* This phase 2 study showed that CCP with high nAb titer (>1:640) provided more rapid recovery to hospital discharge and fewer COVID-19 attributable AEs than CCP with nAb titer between the FDA-recommended minimum standard and 4-fold higher (≥1:160-1:640). The hazard ratio of time to hospital discharge from baseline through day 55, accounting for death as a competing event, contrasting patients receiving high versus standard CCP titer was 1.94 (95% CI 1.05-3.58). Adjusted hazard ratios of high versus standard titer CCP receipt for time to hospital discharge were consistent with the primary unadjusted findings. Mortality through 55 days was lower in the high titer group, but with a wide confidence interval that did not reach statistical significance.

*Implications of all available evidence:* Our data that CCP with nAb >1:640 expedites recovery of patients admitted with COVID-19 compared with CCP with nAb ≥1:160-1:640 suggests that a threshhold of nAb ≥1:160 may be too low to define CCP as ‘high titer’. Analyses in larger CCP trials should consider full reporting of nAb in CCP units administered at individual study participant level, and specifically whether CCP contained nAb >1:640. Further investigation of CCP with nAb >1:640 is warranted given that raising the threshhold of nAb, or a correlative specific anti-spike antibody assay, used to qualify ‘high titer’ CCP in clinical trials could inform policy guidance and clinical use of CCP.

## Introduction

COVID-19 Convalescent Plasma (CCP) was one of the first putative therapies to become widely adopted after the emergence of the novel SARS-CoV-2 virus. Since the start of the pandemic and FDA authorization for inpatient use in September 2020, CCP has been infused to >500,000 hospitalized patients in the United States.^1^ However, data from randomized control trials (RCTs) comparing CCP to standard-of-care, or CCP to plasma devoid of anti-SARS-CoV-2 antibodies remain mixed. Reported benefits in accelerated recovery and decreased mortality in some CCP RCTs,^2^ have not been observed in the largest RCTs.^3-5^ Underlying the challenges in conducting and interpreting CCP clinical trials are heterogeneity in the source of the CCP product and wide inter-individual variability in the breadth and potency of anti-SARS-CoV-2 neutralizing antibodies (nAbs) and antibodies with non-neutralizing functions between different CCP donors.^6^ Indeed, secondary analyses in CONCOR-1 identified that antibody content and/or CCP supplier had a significant effect on estimates of 30 day mortality and the need for ventilation among the CCP-treated participants.^4^ Findings in large multicenter community-based observational studies suggest that CCP with higher antibody levels is associated with a lower risk of in-hospital mortality,^7-9^ and more recently, use of high antibody titer CCP in outpatients reduced risk for hospitalization.^10^ These findings led to emergency use authorization from the FDA and recommendations from expert societies to use high antibody titer CCP as an oupatient therapeutic for high-risk individuals when other therapies are not available.

Antibody-content is considered a major determinant of CCP safety and effectiveness, however, there remains no standardized assay for distinguishing high antibody titer CCP with potential clinical benefit from CCP without benefit or CCP that may be harmful.^4^ For example, the FDA initially recommended neutralizing antibody titers of >1:160 as qualification for CCP as treatment. However, the limited ability to perform live viral neutralization assays prohibited most trials from using this threshold. Rather, large trials like RECOVERY, qualified high titer CCP using an anti-SARS-CoV-2 spike IgG antibody ELISA (EUROIMMUN) with index value of > 6.0.^3^ In contrast, CCP in the US expanded-access program (EAP) observational study in 30,000 patients used the OrthoVitros ELISA platform (OrthoVitros) to quantify anti-SARS-CoV-2 antibody content to assign high-antibody and lower-antibody CCP.^7^ While these spike-protein targeted ELISA correlate with neutralizing antibody titers, we and others have shown that spike protein IgG titers, including IgG titers targeted to the spike protein receptor binding domain (RBD), correlate but alone they are imperfect surrogates for precise *functional* viral neutralizing capability of CCP.^11,12^

To circumvent the limitation of relying on a single antibody-binding assay as a measure of the global anti-viral activity of a polyclonal therapy like CCP, we designed a study administering CCP with pre-defined ranges of neutralizing antibody. Using an on-site SARS-CoV-2-WA1 viral reporter neutralization assay also used in the development of mRNA vaccines, we precisely defined the *functional* viral inhibitory properties of CCP. Beginning in April 2020, our medical center began collecting locally sourced CCP for use in our hospital.^6^ Here we report outcomes from a pilot phase 2 double-blind study in patients admitted with severe COVID-19 using pre-treatment nAb titer-defined CCP from our donor cohort in two different ranges: FDA-minimum recommend nAb titer of 1:160-1:640 (standard titer) and >1:640 (high titer).

## Methods

### Study design and participants

The CoronaVirus Inactivating Plasma (CoVIP) study was designed as a double-blind, randomized phase 2 trial of COVID-19 Convalescent Plasma with defined neutralizing antibody titers at minimum recommended titers (≥1:160) compared with COVID-19 Convalescent Plasma with a 4-fold higher neutralizing antibody titers (>1:640).^13^ CoVIP was conducted at The University of North Carolina at Chapel Hill (UNC) in accordance with FDA IND 22282 (ClinicalTrials.gov registration *NCT04524507*). The original protocol was designed to enroll 56 participants in a 1:1 randomization schema to receive either standard or high titer CCP.

Eligible participants were adults ≥18 years of age hospitalized with SARS-CoV-2 infection defined as laboratory confirmation with a specific SARS-CoV-2 PCR. Additional inclusion criteria included having one or more respiratory or gastrointestinal symptoms including but not limited to: cough, shortness of breath, difficulty breathing, sore throat, loss of taste, loss of smell, diarrhea, nausea or vomiting. To be eligible, onset of symptoms had to be ≤ eight days prior to admission as defined as self-reported fever or documented fever ≥38.0°C. Patients without subjective or objective fever, but other symptoms consistent with COVID-19 were enrolled or excluded by study PI discretion. Key exclusion criteria were ongoing or prior receipt of immune-based therapies including pooled immunoglobulin within the past 30 days, antibody or T-cell based therapies specific to SARS-CoV-2, contraindication to blood transfusion, absence of ABO-compatible plasma, and inability to infuse the first unit of CCP within 48 hours of enrollment.

Participants or Legally Authorized Representatives (LAR) provided written (electronic or paper) informed consents. Separate consent for blood transfusion was also obtained according to institutional standard operating procedures. The clinical trial protocol and informed consent (both in English and Spanish Language) were approved by the Institutional Review Board at UNC-CH (IRB 20-1544). The study was done in accordance with the principles stated in the Declaration of Helsinki and Good Clinical Practice guidelines.

### Intervention

Each participant was to receive two units of CCP (a total of ∼400 – 500 mL) within 48 hours of randomization. The two units could be infused up to 24 hours apart. All eligible participants also received institutional-guided standard-of-care.

### Randomisation and masking

Participants were randomly assigned in a 1:1 ratio to receive either high or standard titer CCP, with randomization stratified by ABO blood group, using permuted block group randomization. During the conduct of the study 13 participants randomized to receive high titer received standard titer CCP because appropriate ABO-compatible high titer CCP was not available. The decision to allow patients access to this therapy was made by the primary investigators and a protocol deviation was approved by the IRB at a time when treatment options were limited and there was equipoise regarding CCP titer. Unmasking was limited to the Blood Bank physician co-investigator who needed access to titer levels to provide appropriate ABO-matched CCP to participants. All study coordinators, clinical physician investigators, treating physicians, participants, and other members of the participants’ health care teams remained masked to treatment assignment. All units of CCP contained a standardized label without designation of neutralizing antibody titer such that healthcare providers, study personnel, and participants remained masked to titer assignment.

We used an honest-broker screening procedure centralized in the Division of Infectious Diseases at UNC-CH for referrals. In this procedure a non-study staff screener filtered new admissions for COVID-19 twice daily and arbitrated eligibility to this and other ongoing intervention studies in the hospital. Study staff were contacted with potential eligible participants. Study staff also interfaced with treating teams to confirm appropriateness to approach for the study.

### Procedures

#### Donors

Convalescent plasma from volunteers who had recovered from SARS-CoV-2 infection was collected by apharesis in the UNC Blood Donation Center (BDC) in accordance with the UNC BDC standard operating procedures and stored on-site. All donations adhered to FDA guidances for CCP, and further details for CCP donor recruitment have been previously described. ^6^ Documentation of SARS-CoV-2 infection by nucleic acid amplification test or antibody test was required to qualify for donation. For measurement of anti-SARS-CoV-2 antibodies, blood from the apheresis diversion pouch was collected and transported on ice for fractionation into serum and cellular components as approved by UNC IRB #20-1141.

#### Anti-SARS-CoV-2 antibody assays

Neutralizing antibody titers were measured using SARS-CoV-2-WA1 viral reporter neutralization assay expressing a nano-luciferase gene, and was recovered using reverese genetics as previously described.^14^ Neutralization assays were performed as previously described. ^6,11,14^ This same assay has been used to assess mRNA vaccine effectiveness.^15^ The minimum threshold for CCP in the standard titer group was set at the initial FDA initial recommended titer of ≥1:160 (standard titer). The high nAb-titer minimum was set to at ≥ 4-fold difference higher than the standard titer group, and to exceed the upper limit of viral neutralization plaque-assay based titers reported at the time of study design (>1:640).^16^

Binding antibody assays to SARS-CoV-2 spike protein receptor binding domain (RBD) and nucleocapsid (N) IgG (Abbott Laboratories) were done as previously described.^6^ Briefly, RBD IgG, IgA and IgM end-point titers were obtained on heat-inactivated serum samples in an in-house developed enzyme-linked immunosorbent assay format starting with a titer of 1:20. ^11^ Nuceocapsid IgG levels were measured in the EUA approved Abbott SARS-CoV-2 IgG assay in a CLIA certified laboratory using the Abbott Architect i2000SR immunoassay analyzer, further described here.^6^

#### Recipients

Participants were monitored by hospital staff during and after each CCP infusion according to standard hospital operating procedures. Clinical symptoms (vital signs and physical exam (as documented by the treating teams), 8-point WHO Ordinal status, laboratory data, and adverse event assessments were obtained on day 1, day 3 (+/- 1), day 7 (+/- 2), day 14 (+/- 2), day 21 (+/- 2), and day 28 (+/- 7). Clinical symptoms were also obtained on participants who elected into an extended study protocol with visits on days 49 (+/- 10), 90 (+/- 14), and 180 (+/- 14). Serum and/or plasma were available from 34 participants (27 in the standard titer group and 7 in the high titer group) prior to CCP administration. To minimize exposures and use of protective equipment by staff, and to minimize sample collection burdens on participants, whenever possible, blood and mucosal lining fluids samples were obtained from remnants of samples collected for clinical purposes.

#### Outcomes

The primary safety endpoint was the cumulative incidence of serious adverse events (SAEs) at study days 14 and 28 after the first CCP infusion. The primary clinical effectiveness endpoint was the days to hospital discharge following the first dose of CCP. Exploratory clinical endpoints included mortality, changes in clinical severity scores (e.g., WHO ordinal clinical status scale), and days of supplemental oxygen, non-invasive ventilation/high-flow oxygen, and/or invasive ventilation/ECMO requirement. All participants were followed until hospital discharge (day 55 post first infusion), with an additional 6 months of follow-up obtained through review of the institutional electronic health record.

### Statistical analysis

Given that 50% of the particpants randomized to the high titer group instead received standard titer CCP, we performed an adjusted as-treated analysis, in effect treating the trial as an observational study accounting for confounding.^17^We used standard statistics to describe patient demographic and clinical characteristics at baseline (date of randomization / first unit of CCP). We compared distributions of baseline characteristics by CCP titer received using Fisher’s exact, Pearson’s chi-square and Kruskal-Wallis tests, as appropriate. We contrasted primary safety endpoints through day 14 and day 28 by CCP titer received using risk differences and 95% confidence intervals as measures of precision. To compare nAb and binding Ab titers by CCP titer received we used the Mann-Whitney-Wilcoxon test.

For the primary clinical endpoint, time from first CCP infusion until hospital discharge, we contrasted the cumulative incidence functions by CCP titer received, accounting for the competing risk of death using Aalen-Johansen estimator and Gray’s test with rho=0. Hazard ratios and 95% confidence intervals for time to hospital discharge by CCP titer were estimated using the Fine-Gray method, accounting for the competing risk of death, with multivariable models fit including only one covariate in each model given the available sample size. Time to death by CCP titer received was compared using the log-rank test and hazard ratios were estimated using Cox proportional hazards models. All hypothesis testing was two-sided. Statistical analyses were performed using SAS software, version 9.4 (SAS Institute) and R software, version 4.0.

### Role of the funding source

The funders of the study had no role in study design, data collection, data analysis, data interpretation, or writing of the report.

## Results

Between August 28 and December 4, 2020, 316 patients were assessed for eligibility and 55 received CCP; 14 and 41 with high and standard titer respectively (**figure 1**). All but one participant received two full units of CCP (54 of 55 patients), with one participant in the standard titer group completing only the first unit. The two units of donor-identical CCP (200-300 mL each) were administered on the day of randomization (baseline), a median of 5.1 hours apart (interquartile range [IQR] 4.4-7.3, full range 0.9-28.7).

**Figure 1.**
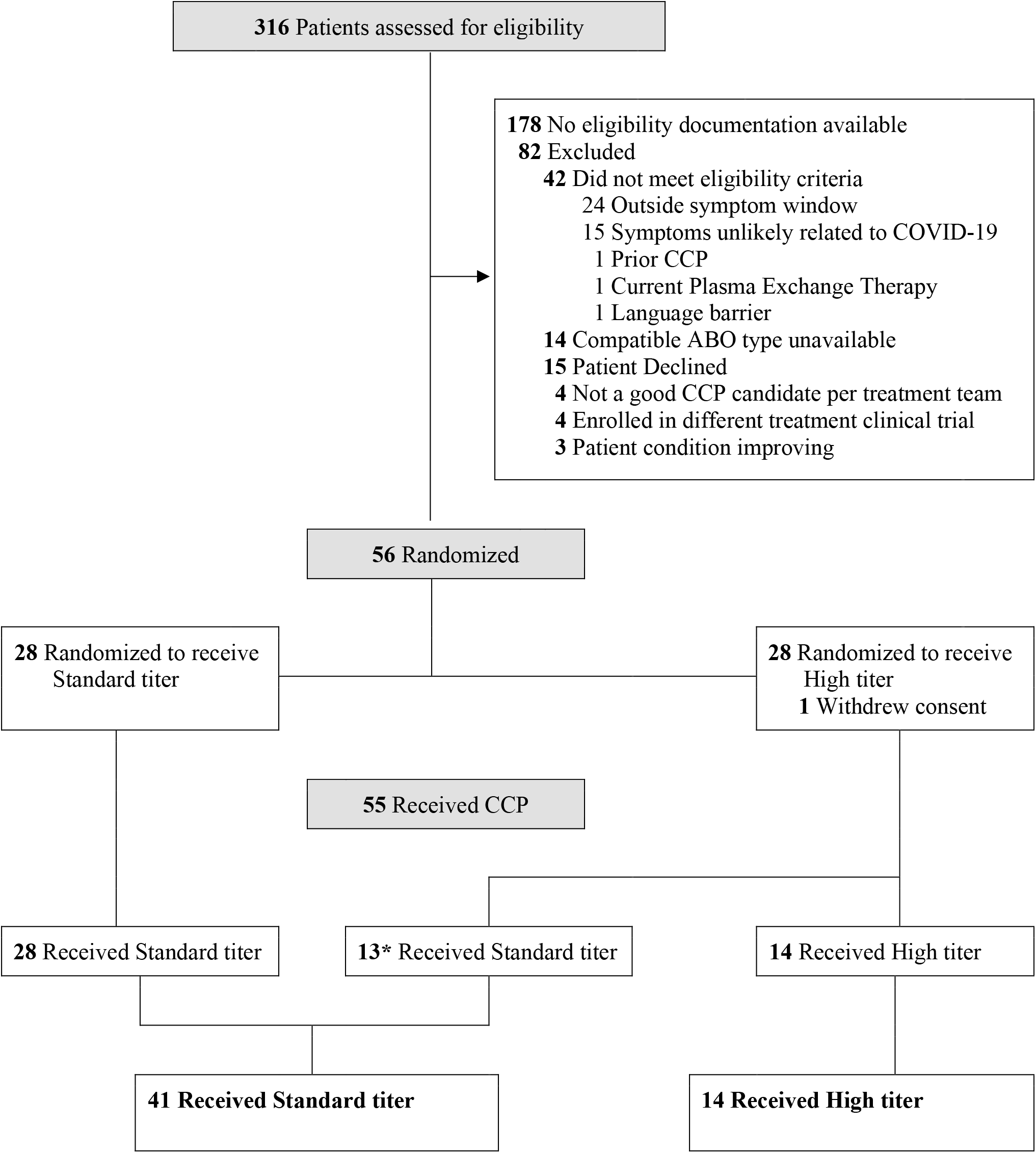
Enrollment, randomization and treatment allocation. Patient flow diagram in the CoVIP study, detailing excluded patients, randomization and CCP titer received. *13 patients randomized to receive high titer CCP received standard titer CPP because ABO-compatible high titer plasma was not available.

Overall patients were a median of 61 years of age (IQR 52-67), 36% female, 25% Black and 33% Hispanic. Demographic and clinical patient characteristics at baseline were comparable by CCP titer received (**table 1**), including respiratory support requirements and the administration other pharmacological COVID-19-directed therapies. Median neutralizing antibody titers from a subset of participant sera prior to CCP infusion were 1:26 (full range undetectable-1:1216) in the high titer group (N=7) and 1:24 (full range undetectable-1:743) in the standard titer group (N=27) (p=0.98).

**Table 1.** Baseline demographic and clinical characteristics.

|  | High Titer<br>(n=14) | Standard Titer<br>(n=41) | p value |
| --- | --- | --- | --- |
| Age, years | 56 (49-75) | 62 (54-67) | 0.68 |
| Female sex | 6 (43) | 14 (34) | 0.75 |
| Race / ethnicity |  |  | 0.50 |
| White | 7 (50) | 14 (34) |  |
| Black | 2 (14) | 12 (29) |  |
| Hispanic | 5 (36) | 13 (32) |  |
| Other | 0 | 2 (5) |  |
| Coexisting Conditions and Medications |  |  |  |
| Any<sup>a</sup> | 13 (93) | 36 (88) | 0.99 |
| Hypertension | 1 (7) | 10 (24) | 0.25 |
| Diabetes | 5 (36) | 19 (46) | 0.54 |
| Obesity | 9 (64) | 23 (56) | 0.76 |
| Cardiovascular disease | 2 (14) | 5 (12) | 0.99 |
| Chronic pulmonary disease | 0 (0) | 9 (22) | 0.09 |
| Solid tumor | 0 (0) | 3 (7) | 0.56 |
| Hematologic malignancies | 0 (0) | 2 (5) | 0.99 |
| Solid organ transplant | 1 (7) | 11 (27) | 0.16 |
| Hematologic stem cell transplantation | 0 (0) | 1 (2) | 0.99 |
| Immunosuppressive medication | 2 (14) | 14 (34) | 0.19 |
| Days from symptom onset | 6 (5-8) | 7 (3-8) | 0.61 |
| Days from hospital admission | 1 (1-2) | 2 (1-2) | 0.47 |
| WHO clinical status<sup>b</sup> | 6 (4-6) | 6 (5-6) | 0.89 |
| Respiratory Support |  |  | 0.63 |
| None | 2 (14) | 6 (15) |  |
| LFNC | 5 (36) | 13 (32) |  |
| Non-invasive ventilation | 5 (36) | 20 (49) |  |
| Invasive ventilation | 2 (14) | 2 (5) |  |
| COVID-19 Medication<sup>c</sup> |  |  |  |
| Remdesivir | 12 (86) | 39 (95) | 0.27 |
| Glucocorticoid | 13 (93) | 37 (90) | 0.99 |
| Anticoagulant | 13 (93) | 39 (95) | 0.99 |
| Antibacterial agent | 7 (50) | 28 (68) | 0.33 |
| Anti-SARS-CoV-2 Neutralizing antibody titer<sup>d</sup> | 1:26 (ND-1:49) | 1:24 (ND-1:76) | 0.98 |

NOTE: ND=not detected; LFNC=low-flow nasal cannula. Data are n (%) or Median (Interquartile Range). P values calculated using Fisher's exact test, Pearson's chi-squared test, or Wilcoxon rank-sum test, as indicated. <sup>a</sup>Any coexisting condition includes any of the conditions listed. <sup>b</sup>WHO ordinal scale with categories: 4= hospitalized, oxygen by mask or nasal prongs, 5=hospitalized, non-invasive ventilation or high-flow oxygen, 6=hospitalized, oxygen by NIV or high flow. <sup>c</sup>COVID-19 medications used at or prior to baseline. <sup>d</sup> Anti-SARS-CoV-2 neutralizing antibody titers available for n=7 of High titer CCP group and n=27 of Standard titer CCP group.

The median nAb titer of CCP units in the high titer group was 1:1080 (IQR 1:827-1:1727, full range 1:667-1:2910) compared to 1:316 (IQR 1:204-1:404, full range 1:161-1:461) in the standard titer group (p<0.0001) (**figure S1**). Additional assays were also performed on the CCP units. The median nAb titer of pseudovirus neutralization assays were higher in the high titer group, 1:1380 (full range 1:113 – 1:29,199) than the standard titer group 1:593 (full range 1:161 – 1:3,455) (p<0.01), but with some overlap between the two groups. Similarly, median anti-RBD IgG end titers were 1:1280 (full range 1:320-1:48,00) in the high titer group and 1:640 (full range 1:40-1:2,520) in the standard titer group (p<0.01). In contrast, there was no difference in the index value for anti-N IgG between the high and standard nAb titer groups (6.36 (range 3.92–6.82) and 5.57 (range 0.72–8.52) respectively (p=0.10)). Spearman’s rank correlation coefficients for nAb and either pseudovirus, anti-RBG IgG, or anti-N IgG were 0.63 (95% CI: 0.42 – 0.77, p< 0.0001), 0.41 (95% CI: 0.15–0.61, p<0.05), and 0.24 (95% CI:-0.03–0.48, p=0.07) respectively.

By day 14 post-infusion, 13 (93%) of the high titer group participants and 37 (90%) of the standard titer group participants experienced at least one adverse event. Most adverse events were categorized as mild (grade 1) or moderate (grade 2) in severity regardless of treatment group. Through 28 days, 27 participants (49%) experienced a grade 3 or greater adverse event. Among these participants, 29% (4 of 14) participants in the high titer group and 56% (23 of 41) participants in the standard titer group had grade 3 or greater adverse events. Cumulative adverse events graded as greater than severe (ie. life-threatening and/or fatal) through 14 days occurred in 0 (0.0%) of 14 high titer participants and 12 (29%) of 41 participants in the standard titer group (p=0.02) (**table 2**). There were no adverse events directly attributable to the CCP infusion in either group. The full list of adverse events through day 28 is presented in **supplementary table 1**.

**Table 2.**
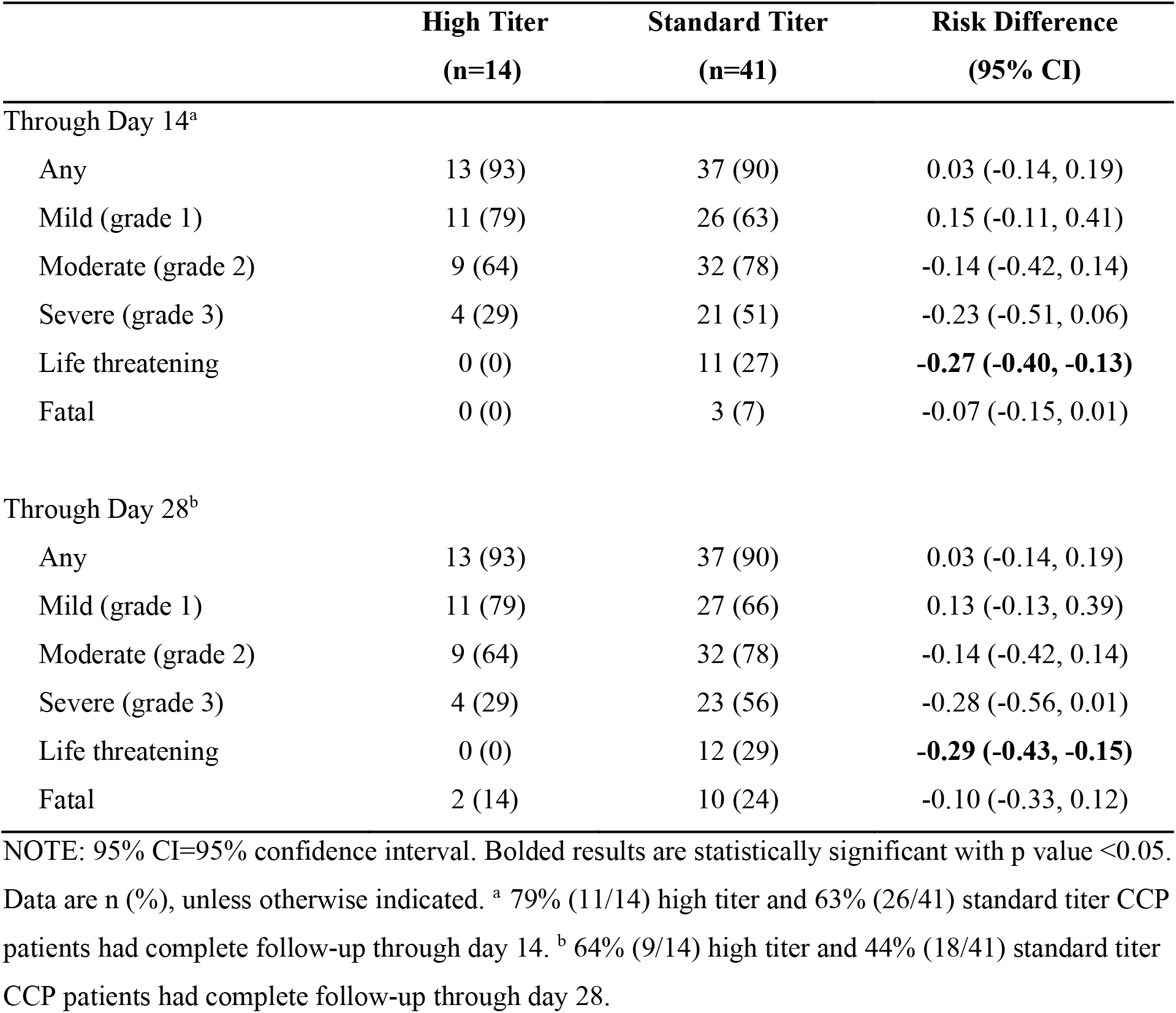
**Primary safety outcomes, stratified by neutralizing antibody titer received and days since first transfusion**

|  | **High Titer**<br>**(n=14)** | **Standard Titer**<br>**(n=41)** | **Risk Difference**<br>**(95% CI)** |
| --- | --- | --- | --- |
| Through Day 14<sup>a</sup> |  |  |  |
| Any | 13 (93) | 37 (90) | 0.03 (-0.14, 0.19) |
| Mild (grade 1) | 11 (79) | 26 (63) | 0.15 (-0.11, 0.41) |
| Moderate (grade 2) | 9 (64) | 32 (78) | -0.14 (-0.42, 0.14) |
| Severe (grade 3) | 4 (29) | 21 (51) | -0.23 (-0.51, 0.06) |
| Life threatening | 0 (0) | 11 (27) | **-0.27 (-0.40, -0.13)** |
| Fatal | 0 (0) | 3 (7) | -0.07 (-0.15, 0.01) |
| Through Day 28<sup>b</sup> |  |  |  |
| Any | 13 (93) | 37 (90) | 0.03 (-0.14, 0.19) |
| Mild (grade 1) | 11 (79) | 27 (66) | 0.13 (-0.13, 0.39) |
| Moderate (grade 2) | 9 (64) | 32 (78) | -0.14 (-0.42, 0.14) |
| Severe (grade 3) | 4 (29) | 23 (56) | -0.28 (-0.56, 0.01) |
| Life threatening | 0 (0) | 12 (29) | **-0.29 (-0.43, -0.15)** |
| Fatal | 2 (14) | 10 (24) | -0.10 (-0.33, 0.12) |

NOTE: 95% CI=95% confidence interval. Bolded results are statistically significant with p value <0.05.
Data are n (%), unless otherwise indicated. <sup>a</sup> 79% (11/14) high titer and 63% (26/41) standard titer CCP patients had complete follow-up through day 14. <sup>b</sup> 64% (9/14) high titer and 44% (18/41) standard titer CCP patients had complete follow-up through day 28.

Patients receiving high versus standard titer CCP had a shorter time to hospital discharge from baseline to day 55, accounting for the competing risk of death, Gray’s p=0.0229 (**figure 2A**). Differences in the cumulative incidence curves for death and hospital discharge favoring patients receiving high versus standard titer CCP were observed across follow-up from baseline through day 55 (**figures 2B and 2C**). The hazard ratio of time to hospital discharge from baseline through day 55 contrasting patients receiving high versus standard CCP titer was 1.94 (95% CI 1.05-3.58) (**figure 3**). Not requiring either non-invasive or invasive ventilation at baseline was strongly associated with a higher rate of hospital discharge (hazard ratio 4.80, 95% CI 2.46-9.36). Other patient characteristics were not associated with hospital discharge by day 55 post CCP infusion in this study population. Adjusted hazard ratios of high versus standard titer CCP receipt on rate of hospital discharge by day 55 post first CCP infusion, were consistent with the primary unadjusted findings (**figure 3**).

**Figure 2.**
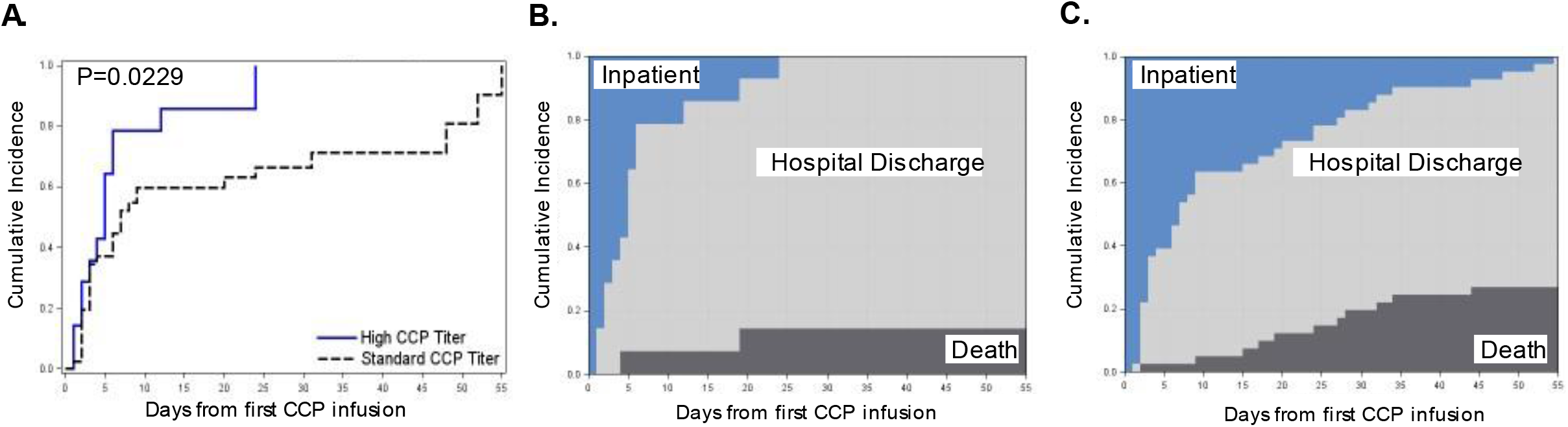
Cumulative incidence of hospital discharge high versus standard neutralizing antibody titer CCP. Time to hospital discharge from first CCP infusion until day 55, by CCP titer received, with death as a competing event, estimated using the Aalen-Johansen estimator, with Gray’s test with rho=0 (Panel A). Stacked cumulative incidence curves for death, hospital discharge and remaining hospitalized, as competing risks, among patients receiving high titer CCP (Panel B), and standard titer CCP (Panel C). Deaths shaded black, hospital discharge gray and remaining hospitalized blue.

**Figure 3.**
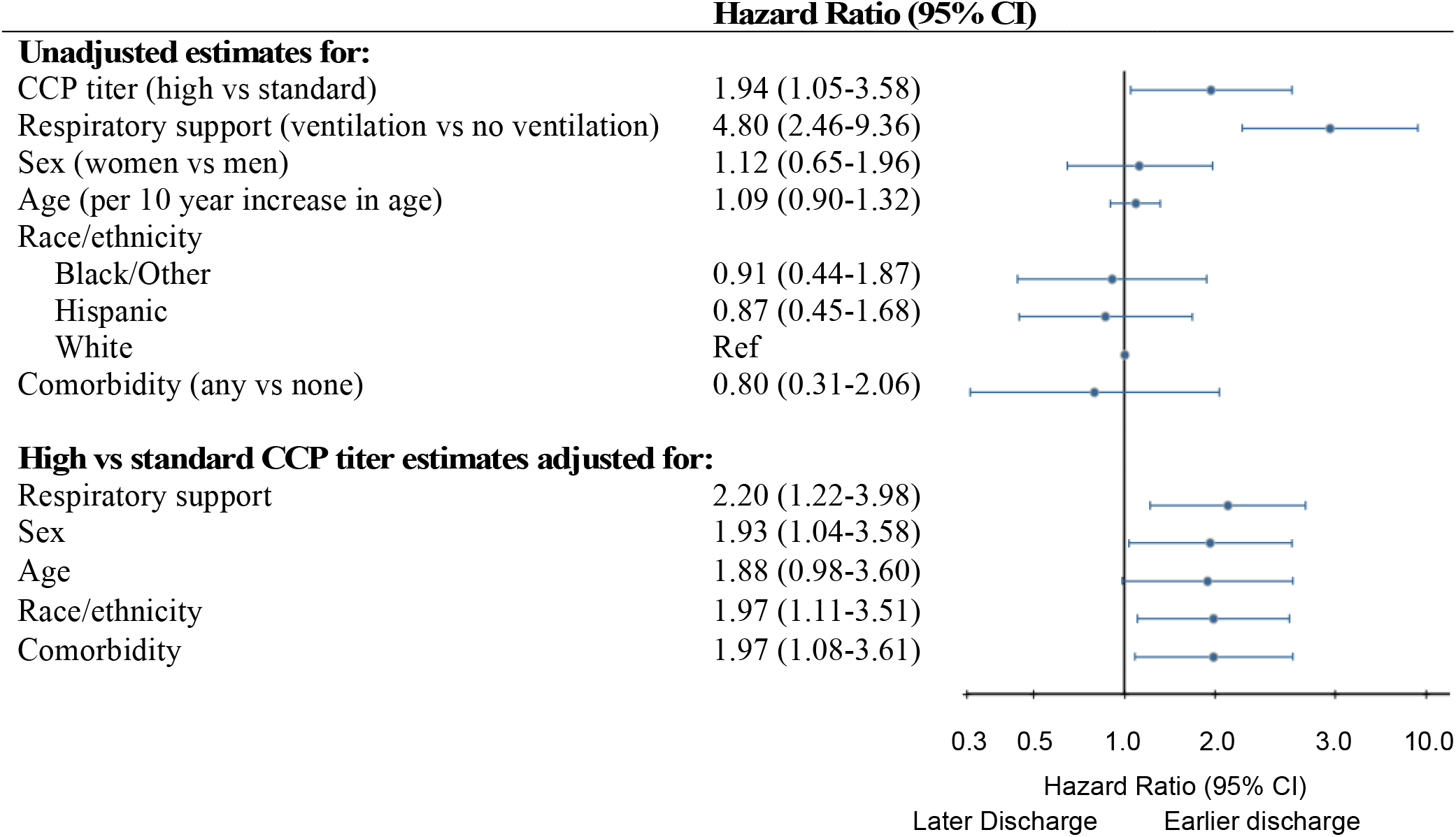
Time to hospital discharge by CCP titer and patient characteristics. Hazard ratios and 95% confidence intervals for hospital discharge from first CCP infusion until day 55, estimated with Fine-Gray method accounting for competing risk of death. Unadjusted estimates for high versus standard CCP titer and select patient characteristics fit with separate models. High versus standard CCP titer adjusted estimates with adjustment made for each patient characteristic in a separate model. Respiratory support measured at baseline contrast is ventilation (i.e., non-invasive or invasive ventilation) versus less than ventilation (i.e., none or low-flow nasal cannula). Comorbidities (any) included diagnosis at baseline of any of the following: hypertension, diabetes, obesity, cardiovascular disease, chronic pulmonary disease, solid tumor, hematologic malignancies, solid organ transplant, and hematologic stem cell transplantation.

In secondary outcome analyses, patients receiving high versus standard titer CCP had a shorter time to hospital discharge from baseline through day 14 and day 28, consistent with day 55 results (**table 3**). Through day 55 post baseline, a greater proportion of patients died in the standard versus high titer CCP group (27 vs 14%, p=0.27), however mortality differences were not statistically significantly different given the imprecision in the estimates. No statistically significant differences in days of respiratory support and WHO clinical status were observed by CCP titer group.

**Table 3.** Secondary outcomes.

|  | High Titer<br>(n=14) | Standard Titer<br>(n=41) | p value |
| --- | --- | --- | --- |
| **Hospital Discharge<sup>a</sup>** |  |  |  |
| Day 14 HR (95% CI) | 1.96 (1.05-3.67) | 1.0 |  |
| Day 28 HR (95% CI) | 2.06 (1.15-3.68) | 1.0 |  |
| **Mortality<sup>b</sup>** |  |  |  |
| Day 14 Deaths, N (%) | 0 (0) | 2 (7) | 0.41<sup>b</sup> |
| Day 14 HR (95% CI) | NA<sup>c</sup> |  |  |
| Day 28 Deaths, N (%) | 2 (14) | 8 (20) | 0.53<sup>b</sup> |
| Day 28 HR (95% CI) | 0.61 (0.13-2.88) | 1.0 |  |
| Day 55 Deaths, N (%) | 2 (14) | 11 (27) | 0.27<sup>b</sup> |
| Day 55 HR (95% CI) | 0.44 (0.10-1.98) | 1.0 |  |
| **Respiratory Support<sup>d</sup>** |  |  |  |
| LFNC days, median (IQR) | 1 (0-3) | 1 (0-3) | 0.57<sup>e</sup> |
| Non-invasive ventilation days, median (IQR) | 1 (0-3) | 1 (0-8) | 0.58<sup>e</sup> |
| Invasive ventilation days, median (IQR) | 0 (0-0) | 0 (0-6) | 0.23<sup>e</sup> |
| Any respiratory support days, median (IQR) | 4 (2-12) | 6 (2-22) | 0.45<sup>e</sup> |
| **WHO clinical status** |  |  | 0.07 / 0.40<sup>e</sup> |
| 0-2 N Day 14 / Day 28 | 12 / 12 | 23 / 25 |  |
| 3 N Day 14 / Day 28 | 0 / 0 | 0 / 3 |  |
| 4 N Day 14 / Day 28 | 0 / 0 | 2 / 2 |  |
| 5 N Day 14 / Day 28 | 1 / 0 | 5 / 0 |  |
| 6 N Day 14 / Day 28 | 0 / 0 | 2 / 0 |  |
| 7 N Day 14 / Day 28 | 1 / 0 | 7 / 1 |  |
| 8 N Day 14 / Day 28 | 0 / 2 | 2 / 10 |  |

NOTE: HR=hazard ratio; 95% CI=95% confidence interval; LFNC=low-flow nasal cannula <sup>a</sup> Hazard ratios and 95% confidence intervals for hospital discharge from first CCP infusion until day 14, 28 and 55, estimated with Fine-Gray method accounting for competing risk of death. <sup>b</sup> Hazard ratios and 95% confidence intervals for time to death from first CCP infusion until day 14, 28 and 55, based on Cox proportional hazards model. P-values based on log-rank test. <sup>c</sup> Hazard ratio not estimable due to number of events. <sup>d</sup> Cumulative number of days of respiratory support through day 55 post first CCP infusion; any respiratory support includes LFNC, non-invasive and invasive ventilation. <sup>e</sup> P-values based on Wilcoxon rank-sum test.

Among 34 patients with available specimens, 21% had a pre-infusion nAb titer > 1:160, with 4, 3, 15 and 12 patients in pre-infusion/CCP nAb titer categories of >1:160/high, >1:160/standard, ≤1:160/high and ≤1:160/standard, respectively (**table 4**). Using the ≤1:160/standard as a reference, there was an interaction between the recipient pre-infusion and CCP donor nAb titer such that the ≥1:160/high group had the greatest estimate of earlier time to discharge (hazard ratio 4.15, 95% CI 1.50-11.46) with the >1:160/standard and ≤1:160/high groups appearing intermediate. Similar trends were seen with mortality hazard ratio estimates, but with wide confidence intervals due to the low sample size.

**Table 4.** **Exploratory clinical outcomes day 55 post-CCP infusion among patients with available pre-infusion anti-SARS-Cov-2 neutralizing antibody (nAb) titer (n=34)**

| CCP nAb titer group | Pre-infusion nAb titer | N | Hospital Discharge<br>HR (95% CI)<sup>a</sup> | Deaths<br>N (%) | Mortality<br>HR (95% CI)<sup>b</sup> |
| --- | --- | --- | --- | --- | --- |
| High; >1:640 | >1:160 | 4 | 4.15 (1.50-11.46) | 0 (0) | NA<sup>c</sup> |
|  | Low (≤1:160) | 15 | 1.64 (0.66-4.10) | 3 (20) | 0.27 (0.06-1.17) |
| Standard; ≥1:160-1:640 | >1:160 | 3 | 1.20 (0.27-5.24) | 1 (33) | 0.56 (0.06-4.84) |
|  | Low (≤1:160) | 12 | 1.0 | 5 (42) | 1.0 |

NOTE: HR=hazard ratio; 95% CI=95% confidence interval; NA=Not available. <sup>a</sup>Hazard ratios and 95% confidence intervals for hospital discharge from first CCP infusion until day 55, estimated using one model with standard nAb titer CCP and low (<1:160) pre-infusion nAb titer as the referent, with Fine-Gray method accounting for competing risk of death. <sup>b</sup> Hazard ratios and 95% confidence intervals for time to death from first CCP infusion until day 55, estimated using one model with low CCP and low pre-infusion titer as the referent, with Cox proportional hazards model. <sup>c</sup> Not estimable due to available sample size.

## Discussion

To our knowledge, this is the first report of the safety and effectiveness of pre-assigned CCP defined as having nAb titer range of 1:160-1:640 versus more than 4-fold higher nAb titer (>1:640). We used these titer range cutoffs based on the initial FDA-guidance that CCP be defined as having nAb ≥1:160, the internal variability of the nAb assay being ∼ 2-fold, and our published findings that the >1:640 represented the top quartile of CCP donors in our program.^6^ We found that patients receiving high titer experienced accelerated time to recovery to hospital discharge, and a trend towards lower mortality through 55 days post-infusion. Adverse events were common in both groups, but were reflective of underlying pathologies attributable to COVID-19 and not from the intervention. Despite a small sample size phase 2 study, unique strengths of our study include pre-defined titer assays assignments using a WT nLuc SARS-CoV-2 viral neutralization assay that is a direct measure of functionally neutralizing antibody and has been used in the development FDA-authorized mRNA-vaccines,^15^ and implementation after corticosteroids and remdesivir had become standard institutional clinical practice and other therapies (like hydroxychloroquine and lopinavir/ritonavir) had fallen out of favor.

Observations that RBD-targeted antibodies have high viral neutralization potential paved the way for rapid development of highly effective vaccines and SARS-CoV-2 directed monoclonal antibody therapies. ^40^ Similarly, we show much stronger correlation between nAb titers and RBD IgG-binding titers than N IgG-index in this cohort. However, overlap exists between RBD IgG-end titers and the two nAb antibody titer groups (**supplementary figure 1**), supporting other findings that a single antibody-binding assay is not fully representive of global anti-viral properties in CCP. Discrepancies in low RBD but high nAb titer CCP may arise from individuals with unmeasured potent nAb targeted at epitopes outside of the RBD, like the N-terminal Domain.^41^ In contrast, high RBD but lower nAb titer CCP could arise from RBD-directed antibodies that poorly compete with SARS-CoV-2 for ACE2 receptor binding. In either case, relying exclusively on a single target antibody ELISA index or end-titer could lead to misclassification of CCP with high neutralizing function, and therefore hinder interpretation of CCP clinical studies.

Significant heterogeneity in clinical therapy studies, such as illness duration and severity, baseline participant charcteristics, and temporal changes in standard of care is not unexpected, especially amidst a rapidly shifting pandemic like Covid-19. However, while most trials target a precisely defining therapeutic dose that is standardized in the intervention arm, the wide inter-individual variability in CCP coupled with a lack of standardized assays for measuring anti-viral and other functional properties precluded this process for CCP. Thus, several of the largest CCP trials used nAb titer-undefined plasma. For example, RECOVERY (n=5795 participants) did not report CCP nAb titer^18^, RE-MAP CAP (n=1075 CC participants) measured nAb titer data on the majority (but not all) of CCP units (median ∼1:160-1:175), but the trial’s focus on critically ill patients at very late stages of Covid-19 (median 43 days after hospital admission) is an outlier beyond the window where any Covid-19 directed therapeutics are now being deployed^19^, and CONCOR-1 enrolled 346 participants in the CCP group, but found significant heterogeneity in clinical outcomes driven by plasma suppliers that also varied in nAb titer ranges.^4^ Among 26 phase 2 or 3 CCP trials identified in PubMed between 2020-2022, 18 measured nAb on all units reported (N=2,692 CCP participants) whereas the other eight either did not perform an end-titer assay^20^, reported nAb titer in only a subset of units^19,21^, or did not report nAb titer^3,22-25^ (N=7,079 CCP partcipants). Using in-study mortality as the most consistently reported primary or secondary outcome, aggregate risk ratios for death in CCP treated-participants versus no CCP comparator groups in trials reporting incomplete or undefined nAb titer (n=8), median nAb < 1:160 (n=7), ^26,27,28,4,29,30,31^ or nAb ≥ 1:160 (n=11)^2,4,5,32-39^ were 0.99 (0.93-1.04), 1.04 (0.86-1.26), and 0.88 (0.73-1.05), respectively (**supplementary figure 2**). The median nAb titer in four studies exceeded 1:320, but only one other study reported a median nAb >1:640 in a CCP intervention arm. Aggregate risk ratios for mortality increasingly favored higher nAb titer CCP compared with no CCP (or lower nAb titer in CoVIP), but with imprecise estimates due to small study size. While larger trials with CCP nAb >1:640 are needed to establish if this very high titer CCP would have clinical benefit, this stratified comparison of CCP studies based on nAb titer infused does raise important need to take caution in interpretation of CCP study outcomes, even among the largest multicenter studies with precise statistical measurements but limited direct measurements of the functional properties of the therapy.

Our findings provide important proof-of-concept that potent neutralizing antibody-based therapies may have a role in the management of patients who rapidly progress to severe COVID-19 and/or miss outpatient therapy windows prior to hospitalization. A post-hoc analysis in RECOVERY showed a benefit of monoclonal antibody therapy among patients hospitalized with COVID-19 who had not yet developed a seroresponse.^18^ In our exploratory analysis we also find that earlier patient seroconversion is associated with better outcome, but in our study the most advantageous outcomes were among those who also received the highest antibody content CCP. These findings may indicate synergy between the nAb in CCP and the recipient pre-infusion nAb. Further investigation is needed to better understand of the potential for polyclonal antibodies to work additively or synergistically together, and whether polyclonal antibodies might decrease the risk of treatment-emergent mutations during RBD-directed monoclonal antibody therapy that can rapidly lead to antibody-resistant variants and subsequently prolonged viral shedding.^42^ In addition, a more complex mixture of neutralizing antibodies, such as CCP from vaccinated individuals after Omicron breakthrough infection, might retain activity against variants of concern like Omicron BA.1 and BA.2. that were not inhibited by several available commercial monoclonal antibodies at the time of their emergence.^43^

Our study has several limitations. First, like several prior CCP RCTs, we had difficulty reaching target enrollment for each of our titer-defined groups. Our inability to identify enough donors with high titer CCP resulted in 50% of the patients randomized to the high nAb titer group instead receiving standard nAb titer CCP. We therefore used a standard observational study analytic approach (ie. adjusted as-treated) considering a conventional intention-to-treat analysis uninterpretable. ^17^ Since treatment assignment deviation was based solely on CCP availability and investigators and treating providers remained blinded to treatment received, we did not observe any confounding and our adjusted analyses were consistent to our crude results. Second, since we could not adequately power a three-arm study, we were unable to include a CCP-free group. Third, we are underpowered in this single-center study to reach a statistical conclusion for outcomes like in-patient mortality. Fourth, since D614G was the most common variant in circulation during out study, we cannot extrapolate our findings to other variants like Omicron BA.1 and BA.2.

As SARS-CoV-2 variants continue to emerge and spread, it is possible that CCP will remain an alternative therapeutic should others be unavailable or ineffective against an emergent variant. We suggest that a definition of high titer CCP exceeding >1:640 gives the greatest confidence in benefit potential, and that future CCP trials deliberately use direct measures of the functional anti-SARS-CoV-2 properties to more precisely pre-assign CCP and avoid infusing low titer CCP that is unlikely to have clinical beneft.

## Data Availability

All data produced in the present study are available upon reasonable request to the authors.

## Acknowledgements

We would like to thank all of our UNC CP donors, the staff at the UNC Blood Donation Center including Hannah Thaxton and Taylor A. Whitaker, and the many volunteers who contributed to this work. This project was supported by the UNC Health Foundation and the North Carolina Policy Collaboratory at the University of North Carolina at Chapel Hill with funding from the North Carolina Coronavirus Relief Fund established and appropriated by the North Carolina General Assembly. The NIH SeroNet Serocenter of Excellence Award, U54 CA260543, supported generation of laboratory data and the following investigators: AJM, LP, SN, SW, DMM, ADS, RB, and LAB. AJM was previously funded by an NIH NIAID T32 AI007151. SN is also funded by the following NIH grants: UNC Center for AIDS Research (P30 AI50410), NA-ACCORD COVID-19 Supplement (U01 AI069918). DRM is funded by an NIH F32 AI152296, a Burroughs Wellcome Fund Postdoctoral Enrichment Program Award, and was previously funded by an NIH NIAID T32 AI007151. HIH is funded by an NIH NIAID T32 AI007001.

## Author Contributions

LAB and DvD conceptualized the study the protocol with input from AJM, HR, SW, YP, RB, AMdS, AL, SN and DvD. LAB, JK, and DMM wrote the study protocol and acquired the IND. LAB and DMM were overall program co-primary investigators. LAB was clinical primary investigator, and HR, AL, DMM, and DvD were clinical co-investigators JK was project administrator. JK, CB, and SN conceptualized the randomization scheme and oversaw randomization coding. SW and YP JK, HIH, LAB and DMM acquired funding. analyzed data, and wrote the orginal draft. AJM, RB, YJH, LP, CC, JLS, RB, and AMdS coneptulatized the laboratory antibody assay methodology, and LAB, AJM, BN, and RB analyzed the laboratory data. CB, JK, and BN curated data. SN validated data. LAB, BN, JK, HIH, and SN performed final analysis. BN made preliminary figures and tables with input from JK, LAB, and final tables and figures were made by SN. LAB wrote the original draft with input from AJM, SN, DvD, and DMM. All authors reviewed and edited the final manuscript. DMM and LAB acquired funding.

## Declaration of Interests

DMM has provided consultancy to Merck outside of this work, and owns common stock in Gilead Sciences. All other authors have no conflicts of interest to declare.

## Data Sharing

De-identified individual participant-level clinical data and antibody assay data will be made available upon request immediately after publication and upon approval of PIs. Study protocol, statistical analysis plan, the clinical study report, and summary outcomes are posted at ClinicalTrials.gov NCT04524507. Analytic code is available upon request and approval of SN.

---

**Supplementary Table 1:** SAEs and AEs through day 28.

|  |  | High Titer |  |  |  | Standard Titer |  |  |  |
| --- | --- | --- | --- | --- | --- | --- | --- | --- | --- |
|  |  | No. of | No. of | No. of |  | No. of | No. of | No. of |  |
| Organ System | Term | Events | Subjects w/ SAE | Subjects at Risk | % with SAE | Events | Subjects w/ SAE | Subjects at Risk | % with SAE |
| Severe adverse event |  |  |  |  |  |  |  |  |  |
| Cardiac disorders | Cardiac arrest | 1 | 1 | 14 | 7% | 0 | 0 | 41 | 0% |
| Gastrointestinal disorders | Gastrointestinal hemorrhage | 0 | 0 | 14 | 0% | 1 | 1 | 41 | 2% |
| Investigations | Alanine aminotransferase increased | 0 | 0 | 14 | 0% | 1 | 1 | 41 | 2% |
| Investigations | Aspartate aminotransferase increased | 0 | 0 | 14 | 0% | 1 | 1 | 41 | 2% |
| Investigations | Blood bilirubin increased | 0 | 0 | 14 | 0% | 1 | 1 | 41 | 2% |
| Investigations | Lymphocyte count decreased | 0 | 0 | 14 | 0% | 2 | 2 | 41 | 5% |
| Investigations | Platelet count decreased | 0 | 0 | 14 | 0% | 1 | 1 | 41 | 2% |
| Metabolism and nutrition disorders | Hyperkalemia | 0 | 0 | 14 | 0% | 1 | 1 | 41 | 2% |
| Respiratory, thoracic and mediastinal disorders | Acute respiratory failure | 1 | 1 | 14 | 7% | 13 | 10 | 41 | 24% |
| Respiratory, thoracic and mediastinal disorders | Acute respiratory distress syndrome | 0 | 0 | 14 | 0% | 4 | 3 | 41 | 7% |
| Respiratory, thoracic and mediastinal disorders | Acute respiratory distress | 0 | 0 | 14 | 0% | 1 | 1 | 41 | 2% |
| Non-severe adverse event |  |  |  |  |  |  |  |  |  |
| Blood and lymphatic system disorders | Anemia | 0 | 0 | 14 | 0% | 18 | 14 | 41 | 34% |
| Blood and lymphatic system disorders | Eosinophilia | 1 | 1 | 14 | 7% | 0 | 0 | 41 | 0% |
| Blood and lymphatic system disorders | Leukopenia | 1 | 1 | 14 | 7% | 3 | 3 | 41 | 7% |
| Blood and lymphatic system disorders | Thrombocytopenia | 0 | 0 | 14 | 0% | 3 | 2 | 41 | 5% |
| Cardiac disorders | Acute myocardial infarction | 1 | 1 | 14 | 7% | 0 | 0 | 41 | 0% |
| Cardiac disorders | Atrial fibrillation | 0 | 0 | 14 | 0% | 4 | 4 | 41 | 10% |
| Cardiac disorders | Bradycardia | 1 | 1 | 14 | 7% | 2 | 2 | 41 | 5% |
| Cardiac disorders | Sinus tachycardia | 1 | 1 | 14 | 7% | 1 | 1 | 41 | 2% |
| Gastrointestinal disorders | Vomiting | 0 | 0 | 14 | 0% | 2 | 2 | 41 | 5% |
| Infections and infestations | Bacteremia | 0 | 0 | 14 | 0% | 3 | 3 | 41 | 7% |
| Infections and infestations | Pneumonia | 1 | 1 | 14 | 7% | 5 | 4 | 41 | 10% |
| Infections and infestations | Pneumonia staphylococcal | 0 | 0 | 14 | 0% | 4 | 3 | 41 | 7% |
| Infections and infestations | Escherichia urinary tract infection | 1 | 1 | 14 | 7% | 0 | 0 | 41 | 0% |
| Infections and infestations | Pneumonia bacterial | 1 | 1 | 14 | 7% | 2 | 2 | 41 | 5% |
| Investigations | Activated partial thromboplastin time prolonged | 1 | 1 | 14 | 7% | 7 | 4 | 41 | 10% |
| Investigations | Alanine aminotransferase increased | 8 | 7 | 14 | 50% | 18 | 14 | 41 | 34% |
| Investigations | Aspartate aminotransferase increased | 5 | 4 | 14 | 29% | 8 | 7 | 41 | 17% |
| Investigations | Blood albumin decreased | 1 | 1 | 14 | 7% | 5 | 5 | 41 | 12% |
| Investigations | Blood bicarbonate decreased | 0 | 0 | 14 | 0% | 3 | 3 | 41 | 7% |
| Investigations | Blood bilirubin increased | 0 | 0 | 14 | 0% | 8 | 6 | 41 | 15% |
| Investigations | Blood creatinine increased | 2 | 2 | 14 | 14% | 4 | 4 | 41 | 10% |
| Investigations | Blood potassium decreased | 1 | 1 | 14 | 7% | 0 | 0 | 41 | 0% |
| Investigations | Blood pressure increased | 0 | 0 | 14 | 0% | 5 | 4 | 41 | 10% |
| Investigations | Hemoglobin decreased | 1 | 1 | 14 | 7% | 0 | 0 | 41 | 0% |
| Investigations | International normalized ratio increased | 0 | 0 | 14 | 0% | 9 | 7 | 41 | 17% |
| Investigations | Lymphocyte count decreased | 2 | 2 | 14 | 14% | 13 | 11 | 41 | 27% |
| Investigations | Neutrophil count decreased | 1 | 1 | 14 | 7% | 2 | 2 | 41 | 5% |
| Investigations | Platelet count decreased | 1 | 1 | 14 | 7% | 3 | 1 | 41 | 2% |
| Investigations | White blood cell count decreased | 0 | 0 | 14 | 0% | 3 | 3 | 41 | 7% |
| Investigations | Ejection fraction decreased | 1 | 1 | 14 | 7% | 0 | 0 | 41 | 0% |
| Investigations | Blood alkaline phosphatase increased | 1 | 1 | 14 | 7% | 4 | 4 | 41 | 10% |
| Metabolism and nutrition disorders | Hyperglycemia | 2 | 2 | 14 | 14% | 7 | 7 | 41 | 17% |
| Metabolism and nutrition disorders | Hyperkalemia | 2 | 1 | 14 | 7% | 11 | 10 | 41 | 24% |
| Metabolism and nutrition disorders | Hypernatremia | 1 | 1 | 14 | 7% | 7 | 7 | 41 | 17% |
| Metabolism and nutrition disorders | Hyperphosphatemia | 0 | 0 | 14 | 0% | 2 | 2 | 41 | 5% |
| Metabolism and nutrition disorders | Hypoalbuminemia | 4 | 3 | 14 | 21% | 13 | 11 | 41 | 27% |
| Metabolism and nutrition disorders | Hypocalcemia | 0 | 0 | 14 | 0% | 2 | 2 | 41 | 5% |
| Metabolism and nutrition disorders | Hypoglycemia | 0 | 0 | 14 | 0% | 4 | 4 | 41 | 10% |
| Metabolism and nutrition disorders | Hypokalemia | 1 | 1 | 14 | 7% | 4 | 4 | 41 | 10% |
| Metabolism and nutrition disorders | Hyponatremia | 4 | 4 | 14 | 29% | 9 | 9 | 41 | 22% |
| Metabolism and nutrition disorders | Hypophosphatemia | 2 | 2 | 14 | 14% | 2 | 2 | 41 | 5% |
| Musculoskeletal and connective tissue disorders | Musculoskeletal chest pain | 1 | 1 | 14 | 7% | 0 | 0 | 41 | 0% |
| Psychiatric disorders | Confusional state | 1 | 1 | 14 | 7% | 0 | 0 | 41 | 0% |
| Psychiatric disorders | Delirium | 2 | 2 | 14 | 14% | 0 | 0 | 41 | 0% |
| Respiratory, thoracic and mediastinal disorders | Hypoxia | 1 | 1 | 14 | 7% | 1 | 1 | 41 | 2% |
| Respiratory, thoracic and mediastinal disorders | Pneumothorax | 1 | 1 | 14 | 7% | 0 | 0 | 41 | 0% |
| Skin and subcutaneous tissue disorders | Decubitus ulcer | 1 | 1 | 14 | 7% | 0 | 0 | 41 | 0% |
| Vascular disorders | Hypertension | 0 | 0 | 14 | 0% | 11 | 7 | 41 | 17% |
| Vascular disorders | Hypotension | 0 | 0 | 14 | 0% | 3 | 3 | 41 | 7% |
| Vascular disorders | Venous occlusion | 1 | 1 | 14 | 7% | 0 | 0 | 41 | 0% |

\*\*\*For Non-SAE, only AE that occurred in at least 5% of either arm was reported. In other words, an AE had to occur in one High Titer participant or two Standard Titer participants to be reported in this table

**Supplementary Table 2:**
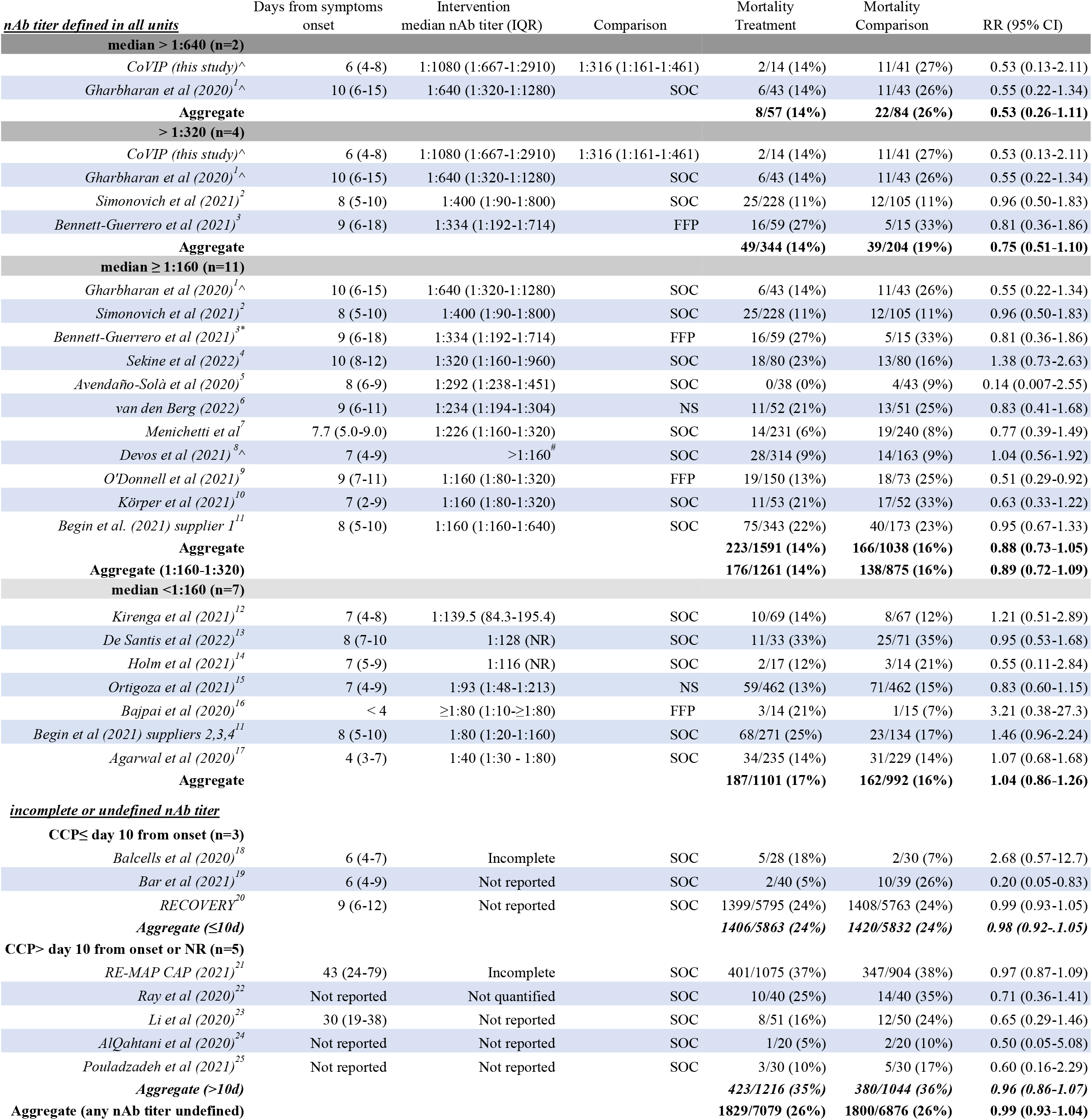
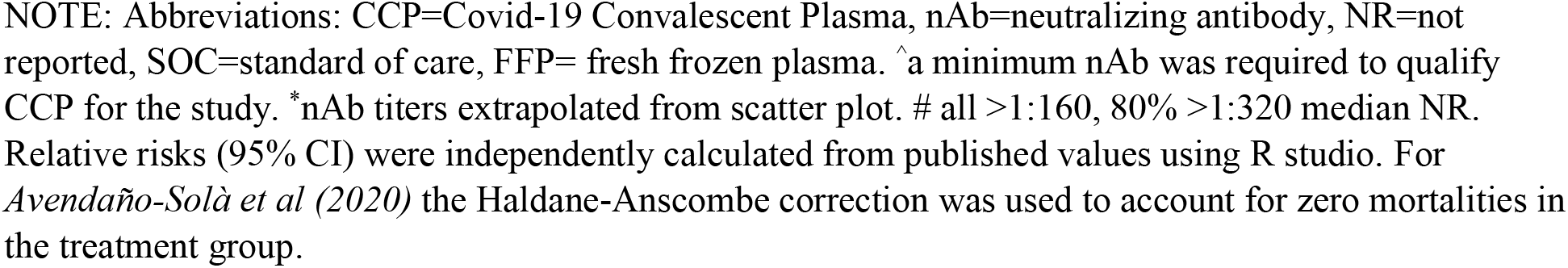
**Mortality (through latest timepoint measured) in randomized control trials of CCP in hospitalized adults organized by neutralizing antibody titer**

| *nAb titer defined in all units* | Days from symptoms<br>onset | Intervention<br>median nAb titer (IQR) | Comparison | Mortality<br>Treatment | Mortality<br>Comparison | RR (95% CI) |
| --- | --- | --- | --- | --- | --- | --- |
| **median > 1:640 (n=2)** |  |  |  |  |  |  |
| *CoVIP (this study)*<sup>^</sup> | 6 (4-8) | 1:1080 (1:667-1:2910) | 1:316 (1:161-1:461) | 2/14 (14%) | 11/41 (27%) | 0.53 (0.13-2.11) |
| *Gharbharan et al (2020)*<sup>1^</sup> | 10 (6-15) | 1:640 (1:320-1:1280) | SOC | 6/43 (14%) | 11/43 (26%) | 0.55 (0.22-1.34) |
| **Aggregate** |  |  |  | **8/57 (14%)** | **22/84 (26%)** | **0.53 (0.26-1.11)** |
| **> 1:320 (n=4)** |  |  |  |  |  |  |
| *CoVIP (this study)*<sup>^</sup> | 6 (4-8) | 1:1080 (1:667-1:2910) | 1:316 (1:161-1:461) | 2/14 (14%) | 11/41 (27%) | 0.53 (0.13-2.11) |
| *Gharbharan et al (2020)*<sup>1^</sup> | 10 (6-15) | 1:640 (1:320-1:1280) | SOC | 6/43 (14%) | 11/43 (26%) | 0.55 (0.22-1.34) |
| *Simonovich et al (2021)*<sup>2</sup> | 8 (5-10) | 1:400 (1:90-1:800) | SOC | 25/228 (11%) | 12/105 (11%) | 0.96 (0.50-1.83) |
| *Bennett-Guerrero et al (2021)*<sup>3</sup> | 9 (6-18) | 1:334 (1:192-1:714) | FFP | 16/59 (27%) | 5/15 (33%) | 0.81 (0.36-1.86) |
| **Aggregate** |  |  |  | **49/344 (14%)** | **39/204 (19%)** | **0.75 (0.51-1.10)** |
| **median ≥ 1:160 (n=11)** |  |  |  |  |  |  |
| *Gharbharan et al (2020)*<sup>1^</sup> | 10 (6-15) | 1:640 (1:320-1:1280) | SOC | 6/43 (14%) | 11/43 (26%) | 0.55 (0.22-1.34) |
| *Simonovich et al (2021)*<sup>2</sup> | 8 (5-10) | 1:400 (1:90-1:800) | SOC | 25/228 (11%) | 12/105 (11%) | 0.96 (0.50-1.83) |
| *Bennett-Guerrero et al (2021)*<sup>3*</sup> | 9 (6-18) | 1:334 (1:192-1:714) | FFP | 16/59 (27%) | 5/15 (33%) | 0.81 (0.36-1.86) |
| *Sekine et al (2022)*<sup>4</sup> | 10 (8-12) | 1:320 (1:160-1:960) | SOC | 18/80 (23%) | 13/80 (16%) | 1.38 (0.73-2.63) |
| *Avendaño-Solà et al (2020)*<sup>5</sup> | 8 (6-9) | 1:292 (1:238-1:451) | SOC | 0/38 (0%) | 4/43 (9%) | 0.14 (0.007-2.55) |
| *van den Berg (2022)*<sup>6</sup> | 9 (6-11) | 1:234 (1:194-1:304) | NS | 11/52 (21%) | 13/51 (25%) | 0.83 (0.41-1.68) |
| *Menichetti et al*<sup>7</sup> | 7.7 (5.0-9.0) | 1:226 (1:160-1:320) | SOC | 14/231 (6%) | 19/240 (8%) | 0.77 (0.39-1.49) |
| *Devos et al (2021)*<sup>8^</sup> | 7 (4-9) | >1:160<sup>#</sup> | SOC | 28/314 (9%) | 14/163 (9%) | 1.04 (0.56-1.92) |
| *O'Donnell et al (2021)*<sup>9</sup> | 9 (7-11) | 1:160 (1:80-1:320) | FFP | 19/150 (13%) | 18/73 (25%) | 0.51 (0.29-0.92) |
| *Körper et al (2021)*<sup>10</sup> | 7 (2-9) | 1:160 (1:80-1:320) | SOC | 11/53 (21%) | 17/52 (33%) | 0.63 (0.33-1.22) |
| *Begin et al. (2021) supplier 1*<sup>11</sup> | 8 (5-10) | 1:160 (1:160-1:640) | SOC | 75/343 (22%) | 40/173 (23%) | 0.95 (0.67-1.33) |
| **Aggregate** |  |  |  | **223/1591 (14%)** | **166/1038 (16%)** | **0.88 (0.73-1.05)** |
| **Aggregate (1:160-1:320)** |  |  |  | **176/1261 (14%)** | **138/875 (16%)** | **0.89 (0.72-1.09)** |
| **median <1:160 (n=7)** |  |  |  |  |  |  |
| *Kirenga et al (2021)*<sup>12</sup> | 7 (4-8) | 1:139.5 (84.3-195.4) | SOC | 10/69 (14%) | 8/67 (12%) | 1.21 (0.51-2.89) |
| *De Santis et al (2022)*<sup>13</sup> | 8 (7-10) | 1:128 (NR) | SOC | 11/33 (33%) | 25/71 (35%) | 0.95 (0.53-1.68) |
| *Holm et al (2021)*<sup>14</sup> | 7 (5-9) | 1:116 (NR) | SOC | 2/17 (12%) | 3/14 (21%) | 0.55 (0.11-2.84) |
| *Ortigoza et al (2021)*<sup>15</sup> | 7 (4-9) | 1:93 (1:48-1:213) | NS | 59/462 (13%) | 71/462 (15%) | 0.83 (0.60-1.15) |
| *Bajpai et al (2020)*<sup>16</sup> | < 4 | ≥1:80 (1:10-≥1:80) | FFP | 3/14 (21%) | 1/15 (7%) | 3.21 (0.38-27.3) |
| *Begin et al (2021) suppliers 2,3,4*<sup>11</sup> | 8 (5-10) | 1:80 (1:20-1:160) | SOC | 68/271 (25%) | 23/134 (17%) | 1.46 (0.96-2.24) |
| *Agarwal et al (2020)*<sup>17</sup> | 4 (3-7) | 1:40 (1:30 - 1:80) | SOC | 34/235 (14%) | 31/229 (14%) | 1.07 (0.68-1.68) |
| **Aggregate** |  |  |  | **187/1101 (17%)** | **162/992 (16%)** | **1.04 (0.86-1.26)** |
| ***incomplete or undefined nAb titer*** |  |  |  |  |  |  |
| **CCP≤ day 10 from onset (n=3)** |  |  |  |  |  |  |
| *Balcells et al (2020)*<sup>18</sup> | 6 (4-7) | Incomplete | SOC | 5/28 (18%) | 2/30 (7%) | 2.68 (0.57-12.7) |
| *Bar et al (2021)*<sup>19</sup> | 6 (4-9) | Not reported | SOC | 2/40 (5%) | 10/39 (26%) | 0.20 (0.05-0.83) |
| *RECOVERY*<sup>20</sup> | 9 (6-12) | Not reported | SOC | 1399/5795 (24%) | 1408/5763 (24%) | 0.99 (0.93-1.05) |
| **Aggregate (≤10d)** |  |  |  | **1406/5863 (24%)** | **1420/5832 (24%)** | **0.98 (0.92-1.05)** |
| **CCP> day 10 from onset or NR (n=5)** |  |  |  |  |  |  |
| *RE-MAP CAP (2021)*<sup>21</sup> | 43 (24-79) | Incomplete | SOC | 401/1075 (37%) | 347/904 (38%) | 0.97 (0.87-1.09) |
| *Ray et al (2020)*<sup>22</sup> | Not reported | Not quantified | SOC | 10/40 (25%) | 14/40 (35%) | 0.71 (0.36-1.41) |
| *Li et al (2020)*<sup>23</sup> | 30 (19-38) | Not reported | SOC | 8/51 (16%) | 12/50 (24%) | 0.65 (0.29-1.46) |
| *AlQahtani et al (2020)*<sup>24</sup> | Not reported | Not reported | SOC | 1/20 (5%) | 2/20 (10%) | 0.50 (0.05-5.08) |
| *Pouladzadeh et al (2021)*<sup>25</sup> | Not reported | Not reported | SOC | 3/30 (10%) | 5/30 (17%) | 0.60 (0.16-2.29) |
| **Aggregate (>10d)** |  |  |  | **423/1216 (35%)** | **380/1044 (36%)** | **0.96 (0.86-1.07)** |
| **Aggregate (any nAb titer undefined)** |  |  |  | **1829/7079 (26%)** | **1800/6876 (26%)** | **0.99 (0.93-1.04)** |

NOTE: Abbreviations: CCP=Covid-19 Convalescent Plasma, nAb=neutralizing antibody, NR=not reported, SOC=standard of care, FFP= fresh frozen plasma. ^a minimum nAb was required to qualify CCP for the study. \*nAb titers extrapolated from scatter plot. # all >1:160, 80% >1:320 median NR. Relative risks (95% CI) were independently calculated from published values using R studio. For *Avendaño-Solà et al (2020)* the Haldane-Anscombe correction was used to account for zero mortalities in the treatment group.

## Unadjusted mortalioty estimates by nAb titer

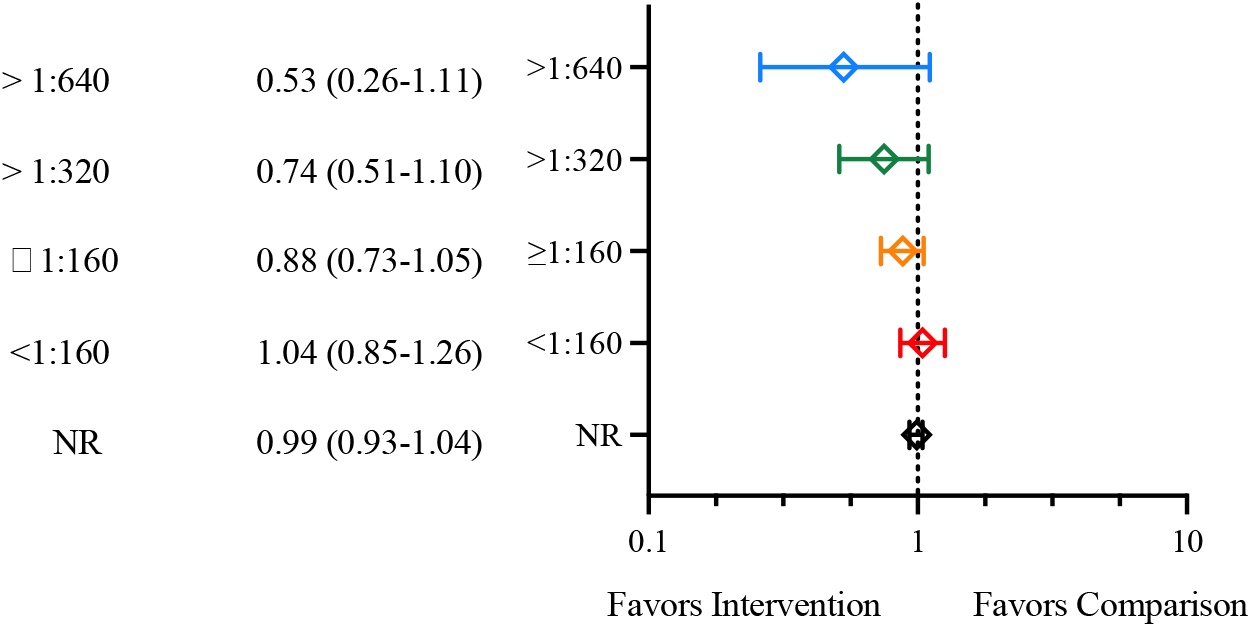

---

**Figure S1.**
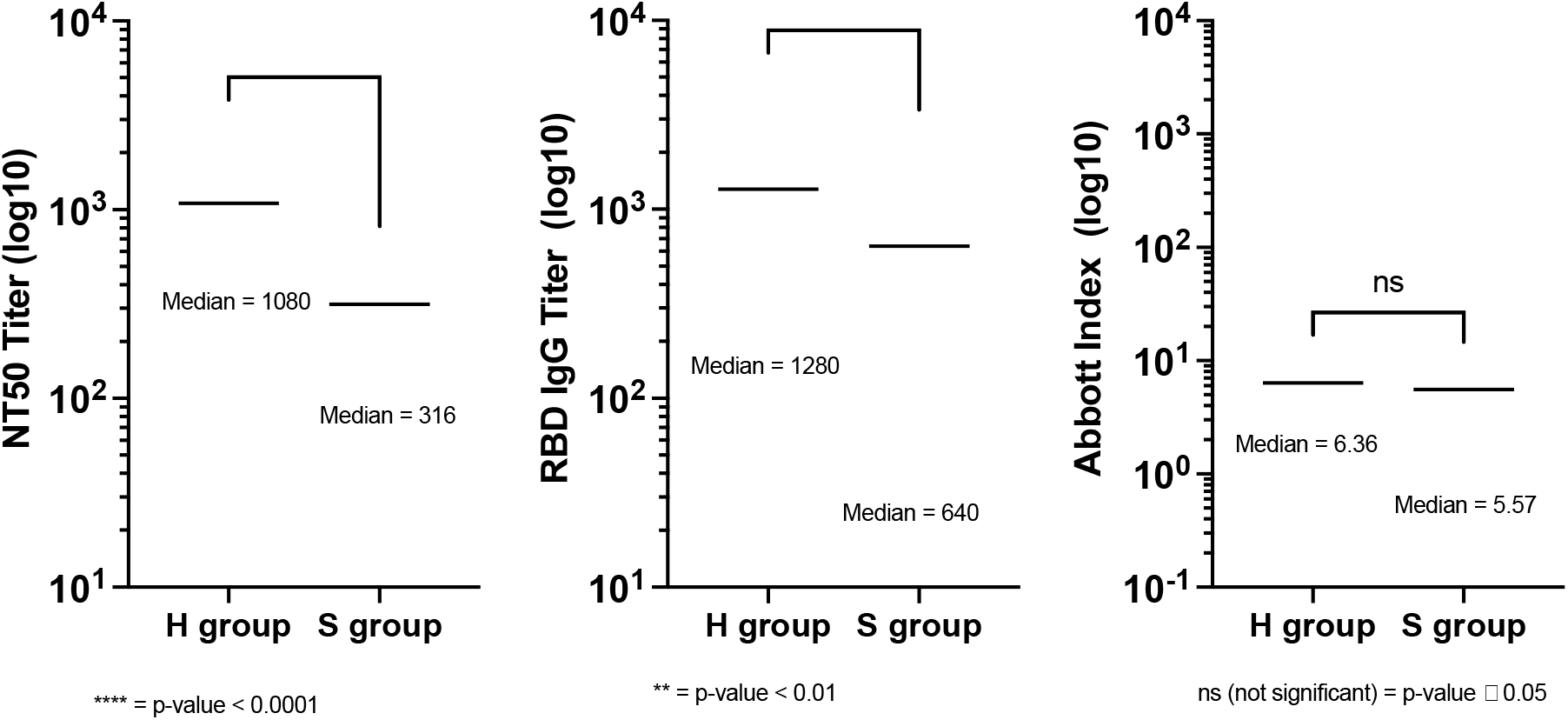
Serological repertoire of CCP units. **anti-SARS-CoV-2 viral neutralization titers and binding antibody ELISAs in CCP units**. A) nAb, B) RBD IgG, and C0 N IgG titer ranges for high titer (H) CCP, and standard titer (S) CCP groups as indicated. Medians shown, analysis done with Mann-Whitney test, p values between H and S groups shown.

## Notes

### Competing Interest Statement

The authors have declared no competing interest.

### Clinical Trial

IND22282
NCT04524507

### Author Declarations

Ethics committee/IRB of the University of North Carolina - Chapel Hill gave ethical approval for this work.

## References

1 Casadevall, A. et al. Convalescent plasma use in the USA was inversely correlated with COVID-19 mortality. Elife 10, doi:10.7554/eLife.69866 (2021).

2 O’Donnell, M. R. et al. A randomized double-blind controlled trial of convalescent plasma in adults with severe COVID-19. J Clin Invest 131, doi:10.1172/JCI150646 (2021).

3 Group, R. C. Convalescent plasma in patients admitted to hospital with COVID-19 (RECOVERY): a randomised controlled, open-label, platform trial. Lancet 397, 2049–2059, doi:10.1016/S0140-6736(21)00897-7 (2021).

4 Begin, P. et al. Convalescent plasma for hospitalized patients with COVID-19: an open-label, randomized controlled trial. Nat Med 27, 2012–2024, doi:10.1038/s41591-021-01488-2 (2021).

5 Avendano-Sola, C. et al. A multicenter randomized open-label clinical trial for convalescent plasma in patients hospitalized with COVID-19 pneumonia. J Clin Invest 131, doi:10.1172/JCI152740 (2021).

6 Markmann, A. J. et al. Sex Disparities and Neutralizing-Antibody Durability to SARS-CoV-2 Infection in Convalescent Individuals. mSphere 6, e0027521, doi:10.1128/mSphere.00275-21 (2021).

7 Joyner, M. J. et al. Convalescent Plasma Antibody Levels and the Risk of Death from Covid-19. N Engl J Med 384, 1015–1027, doi:10.1056/NEJMoa2031893 (2021).

8 Arnold Egloff, S. A. et al. Convalescent plasma associates with reduced mortality and improved clinical trajectory in patients hospitalized with COVID-19. J Clin Invest 131, doi:10.1172/JCI151788 (2021).

9 De Silvestro, G. et al. Outcome of SARS CoV-2 inpatients treated with convalescent plasma: One-year of data from the Veneto region (Italy) Registry. Eur J Intern Med 97, 42–49, doi:10.1016/j.ejim.2021.12.023 (2022).

10 Sullivan, D. J. et al. Early Outpatient Treatment for Covid-19 with Convalescent Plasma. N Engl J Med, doi:10.1056/NEJMoa2119657 (2022).

11 Premkumar, L. et al. The receptor binding domain of the viral spike protein is an immunodominant and highly specific target of antibodies in SARS-CoV-2 patients. Sci Immunol 5, doi:10.1126/sciimmunol.abc8413 (2020).

12 Markmann, A. J. et al. Sex disparities and neutralizing antibody durability to SARS-CoV-2 infection in convalescent individuals. medRxiv, doi:10.1101/2021.02.01.21250493 (2021).

13 Bartelt LA, M. D. Coronavirus Disease 2019 (COVID-19) Antibody Plasma Research Study in Hospitalized Patients (UNC CCP RCT). ClinicalTrials.gov Identifier: NCT04524507.

14 Hou, Y. J. et al. SARS-CoV-2 Reverse Genetics Reveals a Variable Infection Gradient in the Respiratory Tract. Cell 182, 429–446 e414, doi:10.1016/j.cell.2020.05.042 (2020).

15 Anderson, E. J. et al. Safety and Immunogenicity of SARS-CoV-2 mRNA-1273 Vaccine in Older Adults. N Engl J Med 383, 2427–2438, doi:10.1056/NEJMoa2028436 (2020).

16 Shen, C. et al. Treatment of 5 Critically Ill Patients With COVID-19 With Convalescent Plasma. JAMA 323, 1582–1589, doi:10.1001/jama.2020.4783 (2020).

17 Hernan, M. A. & Hernandez-Diaz, S. Beyond the intention-to-treat in comparative effectiveness research. Clin Trials 9, 48–55, doi:10.1177/1740774511420743 (2012).

18 Group, R. C. Casirivimab and imdevimab in patients admitted to hospital with COVID-19 (RECOVERY): a randomised, controlled, open-label, platform trial. Lancet 399, 665–676, doi:10.1016/S0140-6736(22)00163-5 (2022).

19 Writing Committee for the, R.-C. A. P. I. et al. Effect of Convalescent Plasma on Organ Support-Free Days in Critically Ill Patients With COVID-19: A Randomized Clinical Trial. JAMA 326, 1690–1702, doi:10.1001/jama.2021.18178 (2021).

20 Ray, Y. et al. A phase 2 single center open label randomised control trial for convalescent plasma therapy in patients with severe COVID-19. Nat Commun 13, 383, doi:10.1038/s41467-022-28064-7 (2022).

21 Balcells, M. E. et al. Early versus deferred anti-SARS-CoV-2 convalescent plasma in patients admitted for COVID-19: A randomized phase II clinical trial. PLoS Med 18, e1003415, doi:10.1371/journal.pmed.1003415 (2021).

22 Bar, K. J. et al. A randomized controlled study of convalescent plasma for individuals hospitalized with COVID-19 pneumonia. J Clin Invest 131, doi:10.1172/JCI155114 (2021).

23 Li, L. et al. Effect of Convalescent Plasma Therapy on Time to Clinical Improvement in Patients With Severe and Life-threatening COVID-19: A Randomized Clinical Trial. JAMA 324, 460–470, doi:10.1001/jama.2020.10044 (2020).

24 Pouladzadeh, M. et al. A randomized clinical trial evaluating the immunomodulatory effect of convalescent plasma on COVID-19-related cytokine storm. Intern Emerg Med 16, 2181–2191, doi:10.1007/s11739-021-02734-8 (2021).

25 AlQahtani, M. et al. Randomized controlled trial of convalescent plasma therapy against standard therapy in patients with severe COVID-19 disease. Sci Rep 11, 9927, doi:10.1038/s41598-021-89444-5 (2021).

26 De Santis, G. C. et al. High-Dose Convalescent Plasma for Treatment of Severe COVID-19. Emerg Infect Dis 28, 548–555, doi:10.3201/eid2803.212299 (2022).

27 Kirenga, B. et al. Efficacy of convalescent plasma for treatment of COVID-19 in Uganda. BMJ Open Respir Res 8, doi:10.1136/bmjresp-2021-001017 (2021).

28 Holm, K. et al. Convalescence plasma treatment of COVID-19: results from a prematurely terminated randomized controlled open-label study in Southern Sweden. BMC Res Notes 14, 440, doi:10.1186/s13104-021-05847-7 (2021).

29 Bajpai, M. et al. Efficacy of convalescent plasma therapy in the patient with COVID-19: a randomised control trial (COPLA-II trial). BMJ Open 12, e055189, doi:10.1136/bmjopen-2021-055189 (2022).

30 Ortigoza, M. B. et al. Efficacy and Safety of COVID-19 Convalescent Plasma in Hospitalized Patients: A Randomized Clinical Trial. JAMA Intern Med 182, 115–126, doi:10.1001/jamainternmed.2021.6850 (2022).

31 Agarwal, A. et al. Convalescent plasma in the management of moderate covid-19 in adults in India: open label phase II multicentre randomised controlled trial (PLACID Trial). BMJ 371, m3939, doi:10.1136/bmj.m3939 (2020).

32 Gharbharan, A. et al. Effects of Treatment of Coronavirus Disease 2019 With Convalescent Plasma in 25 B-Cell-Depleted Patients. Clin Infect Dis 74, 1271–1274, doi:10.1093/cid/ciab647 (2022).

33 Simonovich, V. A. et al. A Randomized Trial of Convalescent Plasma in Covid-19 Severe Pneumonia. N Engl J Med 384, 619–629, doi:10.1056/NEJMoa2031304 (2021).

34 Bennett-Guerrero, E. et al. Severe Acute Respiratory Syndrome Coronavirus 2 Convalescent Plasma Versus Standard Plasma in Coronavirus Disease 2019 Infected Hospitalized Patients in New York: A Double-Blind Randomized Trial. Crit Care Med 49, 1015–1025, doi:10.1097/CCM.0000000000005066 (2021).

35 Sekine, L. et al. Convalescent plasma for COVID-19 in hospitalised patients: an open-label, randomised clinical trial. Eur Respir J 59, doi:10.1183/13993003.01471-2021 (2022).

36 Van den Berg, K. et al. COVID-19: Convalescent plasma as a potential therapy. S Afr Med J 110, 562–563 (2020).

37 Menichetti, F. et al. Effect of High-Titer Convalescent Plasma on Progression to Severe Respiratory Failure or Death in Hospitalized Patients With COVID-19 Pneumonia: A Randomized Clinical Trial. JAMA Netw Open 4, e2136246, doi:10.1001/jamanetworkopen.2021.36246 (2021).

38 Devos, T. et al. A randomized, multicentre, open-label phase II proof-of-concept trial investigating the clinical efficacy and safety of the addition of convalescent plasma to the standard of care in patients hospitalized with COVID-19: the Donated Antibodies Working against nCoV (DAWn-Plasma) trial. Trials 21, 981, doi:10.1186/s13063-020-04876-0 (2020).

39 Korper, S. et al. Results of the CAPSID randomized trial for high-dose convalescent plasma in patients with severe COVID-19. J Clin Invest 131, doi:10.1172/JCI152264 (2021).

40 Barnes, C. O. et al. SARS-CoV-2 neutralizing antibody structures inform therapeutic strategies. Nature 588, 682–687, doi:10.1038/s41586-020-2852-1 (2020).

41 Voss, W. N. et al. Prevalent, protective, and convergent IgG recognition of SARS-CoV-2 non-RBD spike epitopes. Science 372, 1108–1112, doi:10.1126/science.abg5268 (2021).

42 Rockett, R. et al. Resistance Mutations in SARS-CoV-2 Delta Variant after Sotrovimab Use. N Engl J Med, doi:10.1056/NEJMc2120219 (2022).

43 Li, M. et al. High Viral Specific Antibody Convalescent Plasma Effectively Neutralizes SARS-CoV-2 Variants of Concern. medRxiv, doi:10.1101/2022.03.01.22271662 (2022).

## References

1 Gharbharan, A. et al. Effects of Treatment of Coronavirus Disease 2019 With Convalescent Plasma in 25 B-Cell-Depleted Patients. Clin Infect Dis 74, 1271–1274, doi:10.1093/cid/ciab647 (2022).

2 Simonovich, V. A. et al. A Randomized Trial of Convalescent Plasma in Covid-19 Severe Pneumonia. N Engl J Med 384, 619–629, doi:10.1056/NEJMoa2031304 (2021).

3 Bennett-Guerrero, E. et al. Severe Acute Respiratory Syndrome Coronavirus 2 Convalescent Plasma Versus Standard Plasma in Coronavirus Disease 2019 Infected Hospitalized Patients in New York: A Double-Blind Randomized Trial. Crit Care Med 49, 1015–1025, doi:10.1097/CCM.0000000000005066 (2021).

4 Sekine, L. et al. Convalescent plasma for COVID-19 in hospitalised patients: an open-label, randomised clinical trial. Eur Respir J 59, doi:10.1183/13993003.01471-2021 (2022).

6 van den Berg, K. et al. Convalescent plasma in the treatment of moderate to severe COVID-19 pneumonia: a randomized controlled trial (PROTECT-Patient Trial). Sci Rep 12, 2552, doi:10.1038/s41598-022-06221-8 (2022).

7 Menichetti, F. et al. Effect of High-Titer Convalescent Plasma on Progression to Severe Respiratory Failure or Death in Hospitalized Patients With COVID-19 Pneumonia: A Randomized Clinical Trial. JAMA Netw Open 4, e2136246, doi:10.1001/jamanetworkopen.2021.36246 (2021).

8 Devos, T. et al. A randomized, multicentre, open-label phase II proof-of-concept trial investigating the clinical efficacy and safety of the addition of convalescent plasma to the standard of care in patients hospitalized with COVID-19: the Donated Antibodies Working against nCoV (DAWn-Plasma) trial. Trials 21, 981, doi:10.1186/s13063-020-04876-0 (2020).

9 O’Donnell, M. R. et al. A randomized double-blind controlled trial of convalescent plasma in adults with severe COVID-19. J Clin Invest 131, doi:10.1172/JCI150646 (2021).

10 Korper, S. et al. Results of the CAPSID randomized trial for high-dose convalescent plasma in patients with severe COVID-19. J Clin Invest 131, doi:10.1172/JCI152264 (2021).

11 Begin, P. et al. Convalescent plasma for hospitalized patients with COVID-19: an open-label, randomized controlled trial. Nat Med 27, 2012–2024, doi:10.1038/s41591-021-01488-2 (2021).

12 Kirenga, B. et al. Efficacy of convalescent plasma for treatment of COVID-19 in Uganda. BMJ Open Respir Res 8, doi:10.1136/bmjresp-2021-001017 (2021).

13 De Santis, G. C. et al. High-Dose Convalescent Plasma for Treatment of Severe COVID-19. Emerg Infect Dis 28, 548–555, doi:10.3201/eid2803.212299 (2022).

14 Holm, K. et al. Convalescence plasma treatment of COVID-19: results from a prematurely terminated randomized controlled open-label study in Southern Sweden. BMC Res Notes 14, 440, doi:10.1186/s13104-021-05847-7 (2021).

15 Ortigoza, M. B. et al. Efficacy and Safety of COVID-19 Convalescent Plasma in Hospitalized Patients: A Randomized Clinical Trial. JAMA Intern Med 182, 115–126, doi:10.1001/jamainternmed.2021.6850 (2022).

16 Bajpai, M. et al. Efficacy of convalescent plasma therapy in the patient with COVID-19: a randomised control trial (COPLA-II trial). BMJ Open 12, e055189, doi:10.1136/bmjopen-2021-055189 (2022).

17 Agarwal, A. et al. Convalescent plasma in the management of moderate covid-19 in adults in India: open label phase II multicentre randomised controlled trial (PLACID Trial). BMJ 371, m3939, doi:10.1136/bmj.m3939 (2020).

18 Balcells, M. E. et al. Early versus deferred anti-SARS-CoV-2 convalescent plasma in patients admitted for COVID-19: A randomized phase II clinical trial. PLoS Med 18, e1003415, doi:10.1371/journal.pmed.1003415 (2021).

19 Bar, K. J. et al. A randomized controlled study of convalescent plasma for individuals hospitalized with COVID-19 pneumonia. J Clin Invest 131, doi:10.1172/JCI155114 (2021).

20 Group, R. C. Convalescent plasma in patients admitted to hospital with COVID-19 (RECOVERY): a randomised controlled, open-label, platform trial. Lancet 397, 2049–2059, doi:10.1016/S0140-6736(21)00897-7 (2021).

21 Writing Committee for the, R.-C. A. P. I. et al. Effect of Convalescent Plasma on Organ Support-Free Days in Critically Ill Patients With COVID-19: A Randomized Clinical Trial. JAMA 326, 1690–1702, doi:10.1001/jama.2021.18178 (2021).

22 Ray, Y. et al. A phase 2 single center open label randomised control trial for convalescent plasma therapy in patients with severe COVID-19. Nat Commun 13, 383, doi:10.1038/s41467-022-28064-7 (2022).

24 AlQahtani, M. et al. Randomized controlled trial of convalescent plasma therapy against standard therapy in patients with severe COVID-19 disease. Sci Rep 11, 9927, doi:10.1038/s41598-021-89444-5 (2021).

25 Pouladzadeh, M. et al. A randomized clinical trial evaluating the immunomodulatory effect of convalescent plasma on COVID-19-related cytokine storm. Intern Emerg Med 16, 2181–2191, doi:10.1007/s11739-021-02734-8 (2021).

